# A Global Scale Estimate of Novel Coronavirus (COVID-19) Cases Using Extreme Value Distributions

**DOI:** 10.1101/2020.04.17.20069500

**Authors:** M. Aadhityaa, K. S. Kasiviswanathan, Idhayachandhiran Ilampooranan, B. Soundharajan, M. Balamurugan, Jianxun He

## Abstract

The COVID-19 pandemic has created a global crisis and the governments are fighting rigorously to control the spread by imposing intervention measures and increasing the medical facilities. In order to tackle the crisis effectively we need to know the trajectories of number of the people infected (i.e. confirmed cases). Such information is crucial to government agencies for developing effective preparedness plans and strategies. We used a statistical modeling approach – extreme value distributions (EVDs) for projecting the future confirmed cases on a global scale. Using the 69 days data (from January 22, 2020 to March 30, 2020), the EVDs model predicted the number of confirmed cases from March 31, 2020 to April 9, 2020 (validation period) with an absolute percentage error < 15 % and then projected the number of confirmed cases until the end of June 2020. Also, we have quantified the uncertainty in the future projections due to the delay in reporting of the confirmed cases on a global scale. Based on the projections, we found that total confirmed cases would reach around 11.4 million globally by the end of June 2020.The USA may have 2.9 million number of confirmed cases followed by Spain-1.52 million and Italy-1.28 million.

## Introduction

The first case of respiratory disease, pneumonia, with symptoms similar to the severe acute respiratory syndrome coronavirus (SARS-CoV) was reported in Wuhan City, China in December 2019 [1]. A novel strain of severe acute respiratory syndrome coronavirus 2 (SARS-CoV-2) was confirmed on January 7, 2020 [1, 2]. The novel corona virus (COVID-19), which is the seventh member of the coronavirus family, along with the SARS-CoV and the middle east respiratory syndrome coronavirus (MERS-CoV) spread from animals to humans [2]. Since the reporting of the zero-patient in December 2019, COVID-19 has spread dramatically worldwide and the World Health Organization (WHO) has declared the outbreak as Public Health Emergency of International Concern (PHEIC) on January 30, 2020 [2]. As of April 9, 2020, globally the total number of confirmed, recovered, and mortality cases were 1,595,350, 353,975 and 95,455 respectively [3]. The reported cases globally are drastically increasing from 4,219/day in February 2020 to 50,784/day in early April 2020. The trajectory of the infection spread during the coming days is important information in order to plan, prepare, and scale up the intervention measures including the medical facilities to meet the increased influx of patients and other governing policies to control the transmission.

To date (as of April 9, 2020), the United States of America (USA) has the highest number of affected cases (461,437), followed by Spain (153, 222) and Italy (143,626) [3]. The global case-fatality rate (CFR), which is the ratio of the confirmed deaths to the confirmed cases, has increased from 1.37% on January 19, 2020 to 5.95% by April 9, 2020 [4]. In contrast, the current CFR of China, Japan, and Iran has either reduced or remained constant when compared to their initial CFR [4]. The global CFR is however increasing continuously due to the different timing of the onset of the pandemic in different countries. Overall, the number of confirmed cases has been explosively increasing with time so far.

Modeling tools have been widely used to predict the COVID-19 spread to help the medical professionals, policymakers, and governing bodies for implementing interventions measures to control the pandemic. Since the onset of COVID-19, studies have used different mathematical and dynamic stochastic transmission models to predict the transmission and intervention impacts [5, 6, 7, 8, 9, 10, 11]. However, these epidemiological models involve a number of parameters that are not readily available due to the absence or lack of the data for extracting the knowledge especially during the early period of the outbreak. These parameters have been either assumed or adopted from previous pandemic studies and consequently the performance of these models has been questioned [1, 11].

To address the above shortcomings of epidemiological models, we proposed a statistical modelling approach, using Extreme Value Distributions (EVD) to describe the evolution of the COVID-19 spread and then to generate the future projections. EVDs are used to fit series of observation mainly to estimate extreme events of future that were not observed in the past. Though application of EVDs are very common in earth sciences to model the unusual events, recently, EVDs are applied in health care sector and shown to produce promising results [12].

One of the main challenges in simulating and projecting the COVID-19 spread trajectory is the delay in reporting (R_d_) which is the lag time between onset of symptoms of the disease and date of reporting [13]. The delay in reporting may vary due to various reasons such as delay in (i) reporting at the hospital, (ii) diagnostics, (iii) reporting the confirmed cases in databases etc. Also, R_d_ imposes high uncertainty in estimating the spread trajectory and thus excluding R_d_ in modeling analysis could lead to unrealistic projections with underestimation in the projected confirmed cases [9]. Very few studies have considered R_d_ in their modeling studies to estimate the transmission dynamics of COVID-19 and reported an average R_d_ value of 7.6 days and 6.1 days [9,13].

Therefore, for quantifying the uncertainty in projected cases, we have estimated the fold increase in the confirmed cases due to R_d_. Thus, the key contributions of the study are (i) using EVD theory for the projection of COVID-19 cases, (ii) incorporating R_d_ value to estimate the uncertainty in future projections, and (iii) global scale projection of confirmed and death cases.

## Materials and Method

We collected the daily time series of the number of confirmed and death cases from John Hopkins University Center for Systems Science and Engineering [3] for 177 countries, out of which only 42 countries (refer S1 table) that exceeded 1000 confirmed cases (as on March 30, 2020) were considered for the analysis. These 42 countries spread across different continents except Antarctica and accounts for 96.5% of the total confirmed cases globally as on March 30, 2020. We observed that majority of the countries with significant number of confirmed cases are from Europe and Asia followed by North America and South America.

**S1 Table: Total number of confirmed COVID 19 cases as on March 30, 2020 (List of countries short listed based upon a minimum threshold of 1000 confirmed cases)**

Though the number of COVID-19 infected cases were reported even before January 22, 2020 in China especially in the Hubei province, we have not considered that data in the analysis since the COVID-19 outbreak has been contained and extensive research studies have already been conducted [14, 15, 16, 17].

Application of extreme value distribution has already been explored to model the mortality and morbidity rate associated with pneumonia, influenza and cardiovascular diseases in the public health planning [12, 18]. Therefore, in this paper, we have explored the applicability of EVDs in modeling the confirmed COVID-19 cases. Initial statistical analysis of the data revealed that the critical stage of COVID-19 outbreak largely has no trend in the number of people being infected and therefore the use of EVDs are justified. Among the EVDs, three-parameter distributions such as Generalized Extreme Value (GEV), Generalized Pareto (GP), and Generalized Likelihood (GL) distributions were explored. The parameters of these distributions mainly define the characteristics such as scale, shape and location of the data being fitted using the EVDs. The tail behaviour of the distribution is described by the shape parameter which is estimated from higher order moments, and precise estimation of shape parameter is often computationally difficult [19, 20] and requires suitable moment estimation approaches. Among several methods (i.e. the method of likelihood and the probability weighted moments) for estimating the distribution parameters, the L-moment method has been demonstrated to be more effective in estimating the shape parameters and hence used in this study [21]. As many existing literatures elaborately describe the mathematical description about the extreme value distributions and L-moment methods, the detailed explanations are not provided in this paper.

Conventionally, EVDs have often been applied in the extreme statistical analysis, which estimates the quantities corresponding to specific return periods or probabilities. In this analysis, the sample data from the population are expected to be independent and identically distributed. In the extreme statistical analysis for natural extreme events such as flooding, earthquake, and tsunami, the annual maximum values are often considered. We observed that in the case of COVID-19, the daily recorded confirmed cases are independent with no trend. Thus, the reported confirmed COVID-19 cases were fitted using the EVDs to project the number of future confirmed cases.

In general, the reported cases on any given days were lower than the actual infected cases due to various reasons including the delay in the onset of acute symptoms, inefficiency in the testing methods, lack of sufficient testing facilities etc. There is also significant risk of Covid-19 patients to be tested positive after initially being tested negative due to the inaccuracies in testing and latent symptoms [22]. However, government authorities mainly health care professionals should be aware and be informed about the discrepancy between the reported and actual infected cases to effectively tackle the current COVID-19 situations. Thus, we also estimated the fold increase in the number of confirmed cases due to the delay in reporting. To account the effect of the delay in reporting on the confirmed cases, we considered a Reporting delay (R_d_) of 1 to 7 days and have proposed a simple statistical lagging approach to estimate the fold increase in the number of confirmed cases. For this analysis, the ratio 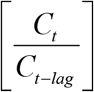 of confirmed cases on the current date *C*_*t*_ to the previously lagged date *C*_*t-lag*_ were computed using the complete data having n data points. The mean value of the fold increase was estimated from the ratios (For example, lag of 5 days will have n-5 number of ratios) for each R_d_. Furthermore, the estimate of the fold increase (with its uncertainty) was coupled with the future projection of the confirmed cases for quantifying associated uncertainty in the projection. Note that other than R_d_, no other sources of uncertainty such as incubation period, communal spread, the effects of lockdown or other containment strategies, healthcare capacity, etc. were included in our study.

## Results and discussion

### Statistical tests and model performance

The daily number of the confirmed cases from the selected 42 countries were computed from the reported cumulative data. These data were further processed with the modified Mann-Kendall test to check for the presence of non-parametric trend and we found that there is no trend in the entire dataset. This proves that the data are statistically independent and identically distributed during the critical stage of pandemic situation. Hence, we applied extreme value distribution to model the available data as well as to project the COVID-19 cases.

As mentioned earlier, three different EVDs were explored to fit the datasets for the numbers of confirmed cases. The root mean squared error (RMSE) computed for each distribution against observation for all datasets are plotted in the boxplot (Fig 1). It is evident from Fig 1 that all the three distributions (GEV, GP and GL) performed equivalently. Furthermore, the fitting performance was slightly improved when using the GP distribution compared to GEV and GL distributions and in particular the GP performed consistently well across all the datasets. In addition, large variations in the estimated distribution parameters (i.e., location, scale, and shape parameters) were identified. The results of parameter variations of GP have been shown in S2 Fig. These variations would reflect the variations in the statistical characteristics of the datasets of different countries. As these models are data specific with parameters not having direct physical meaning, it is hard to link the behaviour of parameters with the modelled variables. Since we observed better performance and lower RMSE using GP distribution, in this study, we are projecting the estimates of confirmed cases for selected countries using the GP models.

**Fig 1.**
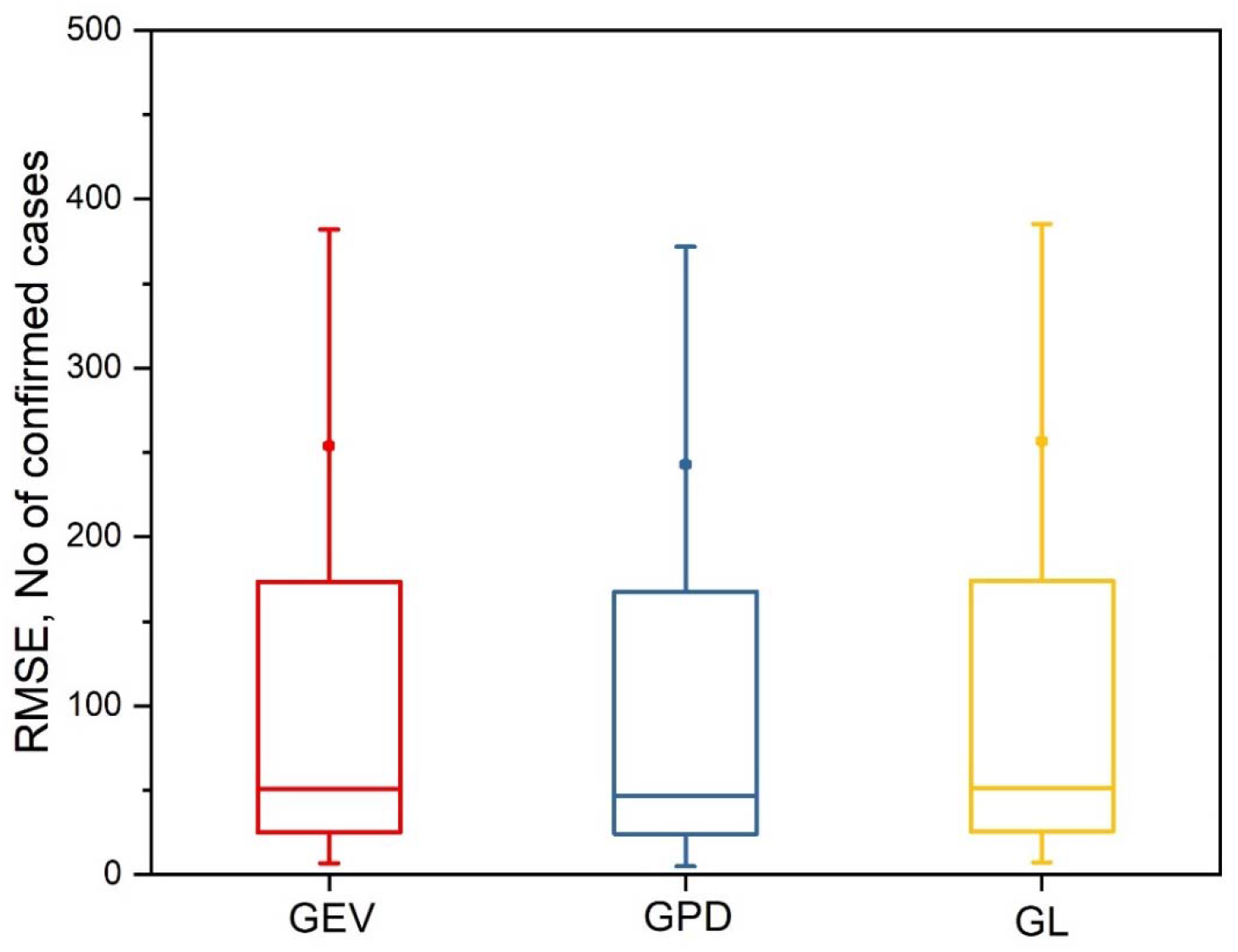
Performance of three different Extreme value probability distributions for fitting the confirmed cases of all 42 countries.

**S2 Fig: Estimated parameters of the GP Models**

Although the outbreak had started in the beginning of January 2020 in Wuhan, China, majority of the countries started experiencing new cases in the beginning of March 2020. All these demonstrates the different timing of the onset of the spread in different countries. However, we considered same period (January 22, 2020 to March 30, 2020) for the model calibration which in turn resulted a uniform data length of all the selected countries for estimating the parameters of distribution.

The model performance has been validated by comparing the model’s projections and the confirmed cases observed for the selected countries for the period of 10 days (March 31 – April 9, 2020). The mean and standard deviation of the resulting percentage error (i.e. ratio of difference between observed minus projected to observed cases) has been shown in Fig 2. Please note that the positive and negative value of mean of the percentage error indicates under and overestimate of the projected value, respectively. From Fig 2, it is evident that the mean and standard deviation of the percentage error are within the ±5% and ±10% respectively for most of the countries, and the overall performance of model is quite satisfactory for majority of the cases except few. The poor performance of model for few countries, for example Germany, Australia, and Iran, might be due to high variation in the infected cases. As more data is available in future, more critical validation of these models can be performed to bring additional insights on the reliability of the model projection.

**Fig 2.**
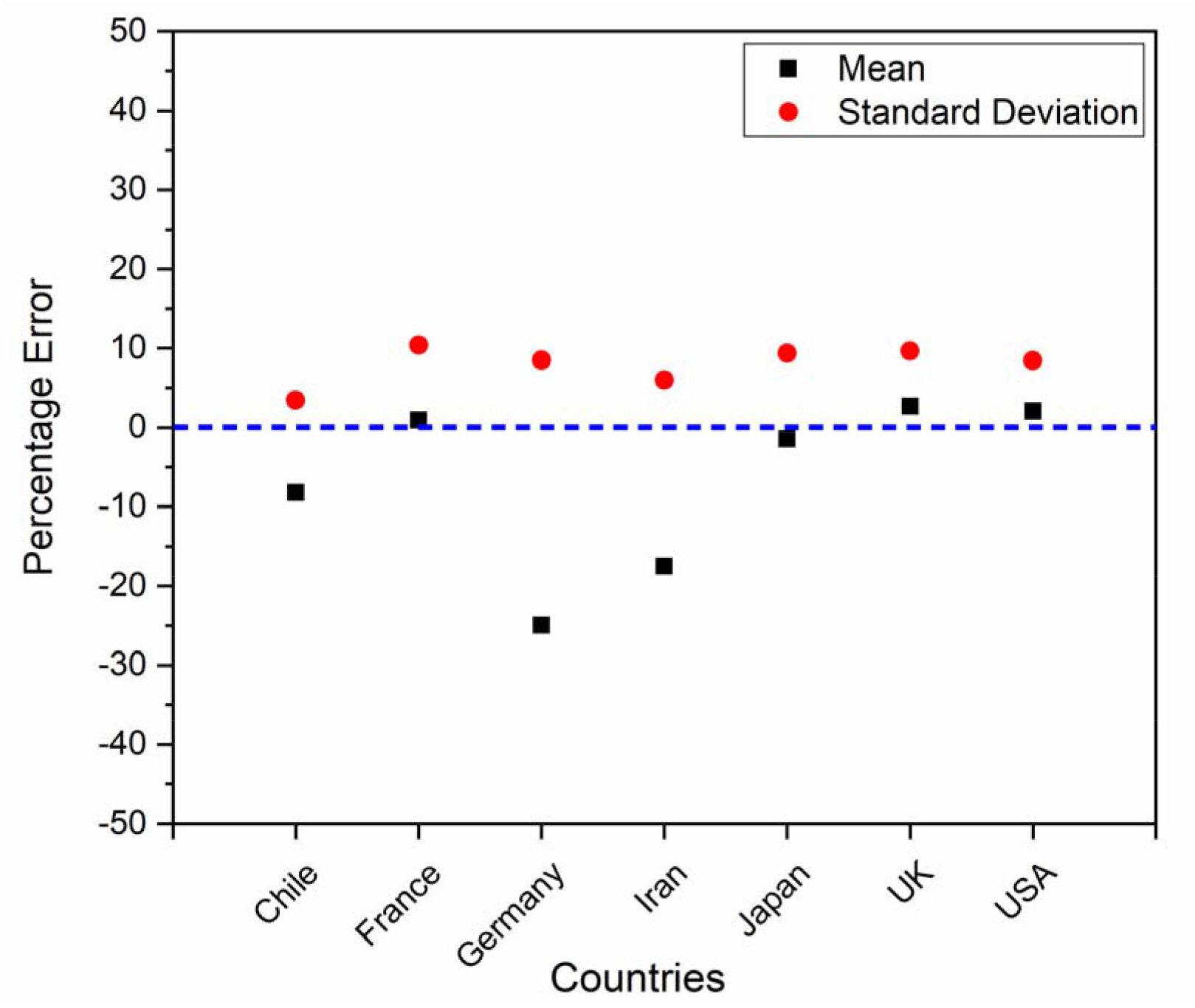
Percentage error in the projected confirmed cases for the validation period from March 31, 2020 to April 09, 2020.

Further, the model was also validated using the projection at global scale. As shown in Table 1, the projections of the confirmed cases are very close to actual value with slight over estimation and the increasing trend is captured well. As mentioned earlier, more data is required to validate the global scale long term projection of model. However, the long-term projection of deaths estimated using these model projections are very close to the projection reported by IHME for the countries such as United States of America (USA) and United Kingdom (UK) [23].

**Table 1.** Global scale estimates of confirmed cases for validation period.

| Date | Projected | Actual |
| --- | --- | --- |
| March 31, 2020 | 869,406 | 857,487 |
| April 1, 2020 | 957,136 | 932,605 |
| April 2, 2020 | 1,045,551 | 1,013,320 |
| April 3, 2020 | 1,134,644 | 1,095,917 |
| April 4, 2020 | 1,224,412 | 1,197,405 |
| April 5, 2020 | 1,314,850 | 1,272,115 |
| April 6, 2020 | 1,405,953 | 1,345,101 |
| April 7, 2020 | 1,497,716 | 1,426,096 |
| April 8, 2020 | 1,590,135 | 1,511,104 |
| April 9, 2020 | 1,683,203 | 1,595,350 |

### Projection of COVID-19 confirmed cases

We observed two main behavioural changes in the number of confirmed cases curves (Fig 3). (i) plateau-peak shift: number of confirmed cases in the USA, Australia, and Italy has plateaued (with approximately 10 cases) soon after the beginning until last week of February 2020 and increased to peak thereafter. A possible reason could be an inflow of passengers soon after the Chinese New Year [24]. (ii) cross-over during March 2020: though the number of confirmed cases around end of February 2020 (approximately 10 cases) was almost similar for the USA, Australia, Italy, and Iran, Australia’s number of confirmed cases were less when compared with the USA, Italy, and Iran during March 2020. The USA’s number of confirmed cases, though lesser than Italy and Iran during the first three weeks of March 2020, has surpassed the cases of Italy and Iran during the end of third week of March 2020 and stayed at peak thereafter. The possible explanation for this behaviour could be attributed to the difference in intervention measures imposed in the respective countries. However, it is important to note that, testing capacity might be one of the keys which dictate the success of non-medical measures such as social-distancing and lockdown to contain the virus [25].

**Fig 3.**
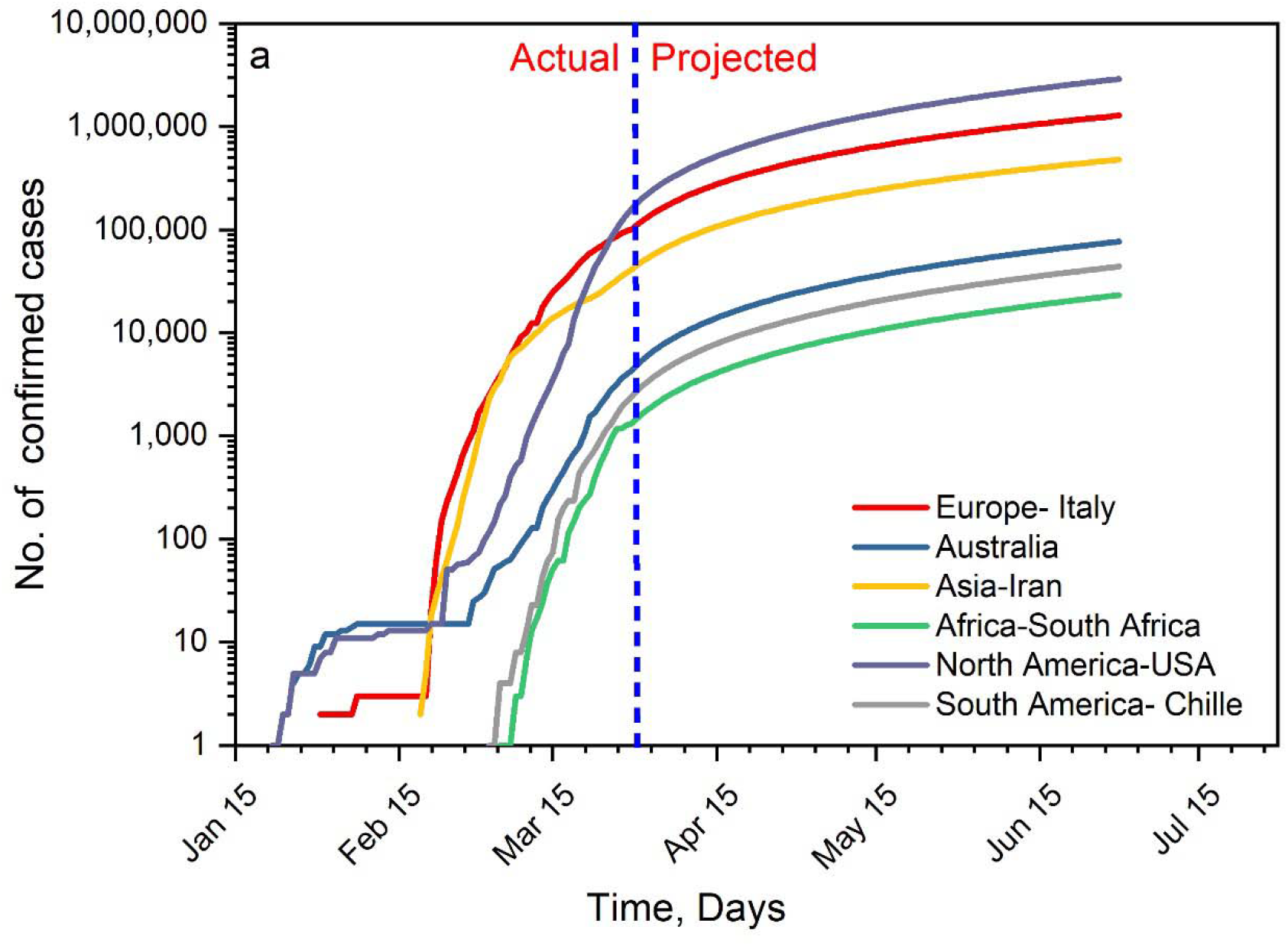
Actual and projected trend of cumulative increase of confirmed cases for the selected countries.

Note that due to the limited observed data for fitting the model, the error in the future projection is expected to be high and increases with time. Thus, the projection was made till June 30, 2020 (three months from March 30, 2020). Along with the estimate of probability, the shape, scale and location parameters of GP distribution were used to project the confirmed cases for the future period. Fig 3 shows the estimate of the number of confirmed cases for the present and future (up to June 30, 2020) for the most affected country of each continent. In Chile and South Africa, during the first week of March 2020, fewer (i.e. around 10 cases) confirmed cases were reported; then an exponential increase was clearly seen during the subsequent weeks. Although number of daily confirmed cases show randomness, the varying pattern across different countries were similar.

As of June 30, 2020, the numbers of the confirmed cases for Italy, Iran, Australia, South Africa and Chile would reach around 1,281,708, 479,531, 76,795, 23,281, and 44,041 respectively. The number of the confirmed cases for the USA would likely to be at least more than one million (highest among all the countries) in the early May 2020, though the initial progression of the number infected was much lower than other highly affected countries such as Italy and Iran. Similar behaviour in the number of projected confirmed cases was observed for the other less affected countries such as India (refer S3 Fig), however, with less magnitude mainly because of delay in onset of disease spread (early March 2020) and reporting of cases. Majority of the countries (mostly developing and under-developed) at the onset of surging increase in COVID-19 spread would neither be equipped with testing facilities nor have the medical infrastructure to tackle the crisis [26]. Therefore, the availability of more data in the forthcoming days would help in producing a more reliable projection of the confirmed cases in the future.

**S3 Fig: Projection of confirmed cases for the countries with delayed onset of COVID-19**

Note that these projections did not explicitly include the effect of the actual stringent control measures (eg. social distancing, travel bans, isolation/quarantine, lockdown) adopted in the various countries at local/regional and national level. However these projections might vary and the number of actual confirmed cases could be less if all the countries apply inter and intra circuit breaking measures (control measures) to reduce the COVID-19 spread. From the projection across the world on June 30, 2020 as illustrated in Fig. 4, 17 thousand (17k) to 3000 thousand (3000k) number of confirmed cases were observed across different countries since January 22, 2020. The maximum number of confirmed cases between 1522k and 2906k were observed in the USA and Spain. Following that Italy and Germany are likely to have more than 1 million confirmed cases.

**Fig 4.**
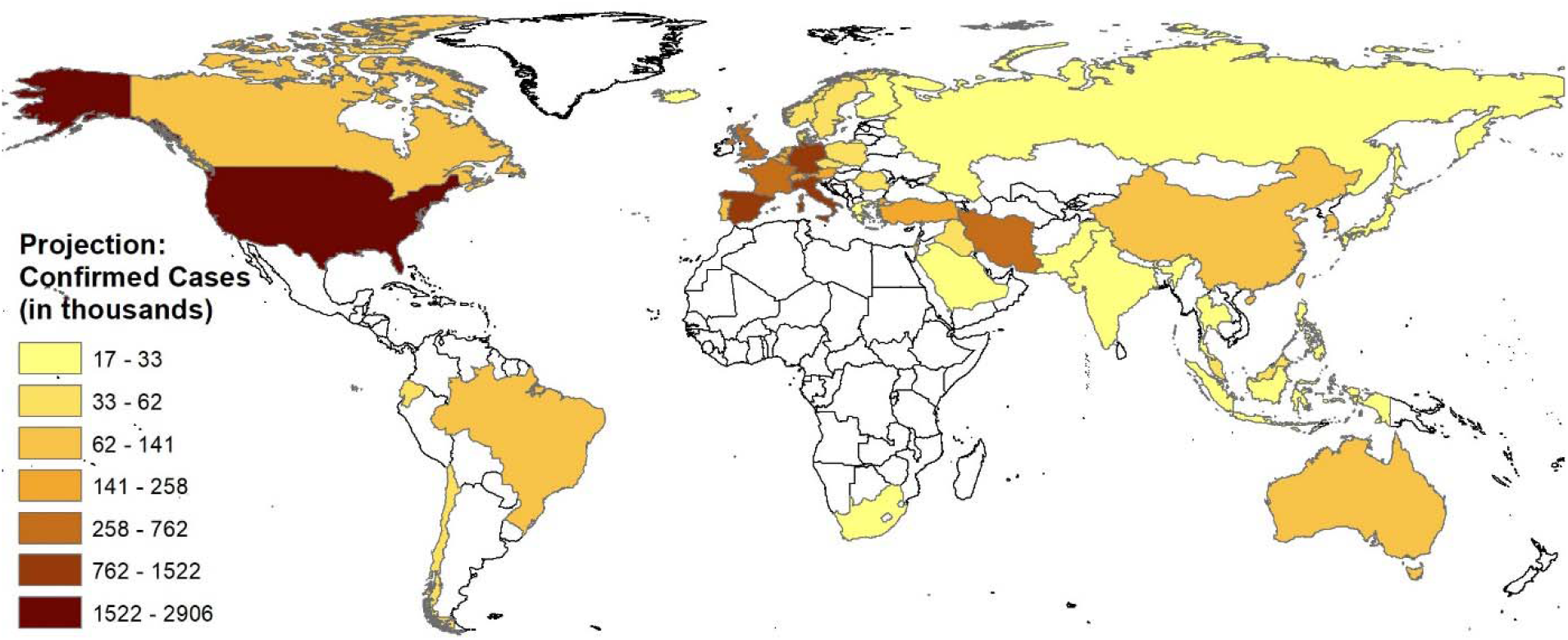
Number of confirmed cases as on June 30, 2020.

Globally the total number of confirmed cases would reach 11.4 million by the end of June 2020. Several countries will exceed one million COVID-19 infections within the next two months. For example, USA with the highest number of confirmed cases globally will be the first to reach the one million count in the first week of May, followed by Spain and Italy in the first week of June 2020. We also estimated that Germany and France will also exceed a million cases in the middle and end of June respectively. Countries like Iran, UK, Chile and Portugal are also identified to be at high risk since confirmed cases in these countries will exceed half a million by the end of June 2020.

The current number of the confirmed cases in India is still in the range of few thousands and the future projection is estimated to be around 28,028, which is considerably lesser than the USA and the UK. However currently in India, stringent measures such as 42 days nation-wide lockdown has been imposed to control and stabilize the communal spread in order to prevent from becoming a global hotspot of COVID-19. It is difficult to project realistic estimates for countries like India and Indonesia due to lack of sufficient data which is attributed to the delay in onset of COVID-19 (first week of March 2020). However, upon getting more data with time, our model could be used to project more realistic values.

In other countries including Japan and South Korea that were severely affected but imposed many preventive measures, would likely to have 23k and 118k confirmed cases respectively. However, the number of confirmed cases might still be reduced depending on the effectiveness of the preventive measures. Please refer S4 Table for the country-wise estimates of projected confirmed cases.

**S4 Table: Country wise estimates of projected confirmed cases**

The rate of increase in the projected confirmed cases were estimated at the end of each months (i.e. April, May and June 2020) including the actual confirmed case (March). This daily rate of confirmed cases was computed from estimating the difference between the cumulative confirmed cases at each month interval and dividing by the total number of days (one month in this case) using the projection of 42 countries. The rate of increase falling in the box (i.e. 25 to 75 percentiles) indicates that in several countries, the impact would be less as the number of confirmed cases per day ranges between few thousand for the projection period varying from April 2020 to June 2020. However, a very high rate of increase in confirmed cases were found in USA, Spain, Italy and Germany followed by France, Iran and UK.

Besides the projection of confirmed cases, we also estimated the likely number of deaths. CFR is commonly used to estimate the risk of death due to any infectious disease. Please note that though CFR (usually represented as percentage of the ratio of confirmed cases to confirmed deaths) is not constant and changes with the context (e.g., it can vary with time, age and the characteristics of infected population, etc.)[4], it can give an approximate estimate of the number of deaths [4, 27, 28, 29].

We estimated the average value of the CFR for the selected 42 countries (from March 15, 2020 to March 30, 2020) and found the CFR values as 9%, 8%, 7%, 6%, 5% and 2% for the countries Italy, Indonesia, Iran, Spain, UK, France and USA respectively. Based on these CFR estimates along with the model projection for the confirmed cases (Fig 6), we estimated the likely number of deaths on June 30, 2020 and found the number of deaths to be highest in Italy (115,354) and Spain (91,340). The number of deaths in countries such as USA, Iran, France and UK are also likely to be high with 58,110, 33,567, 30,490 and 20,116 deaths respectively by the end of June 2020. Refer S5 Table5 for the estimated number of deaths of other countries having CFR greater than 1 percent. Our estimate especially for USA (82,638 deaths by the end of July 2020) is very close with the number of deaths projected by the IHME health service utilization forecasting team (81,114 deaths) [12].

**Fig 5.**
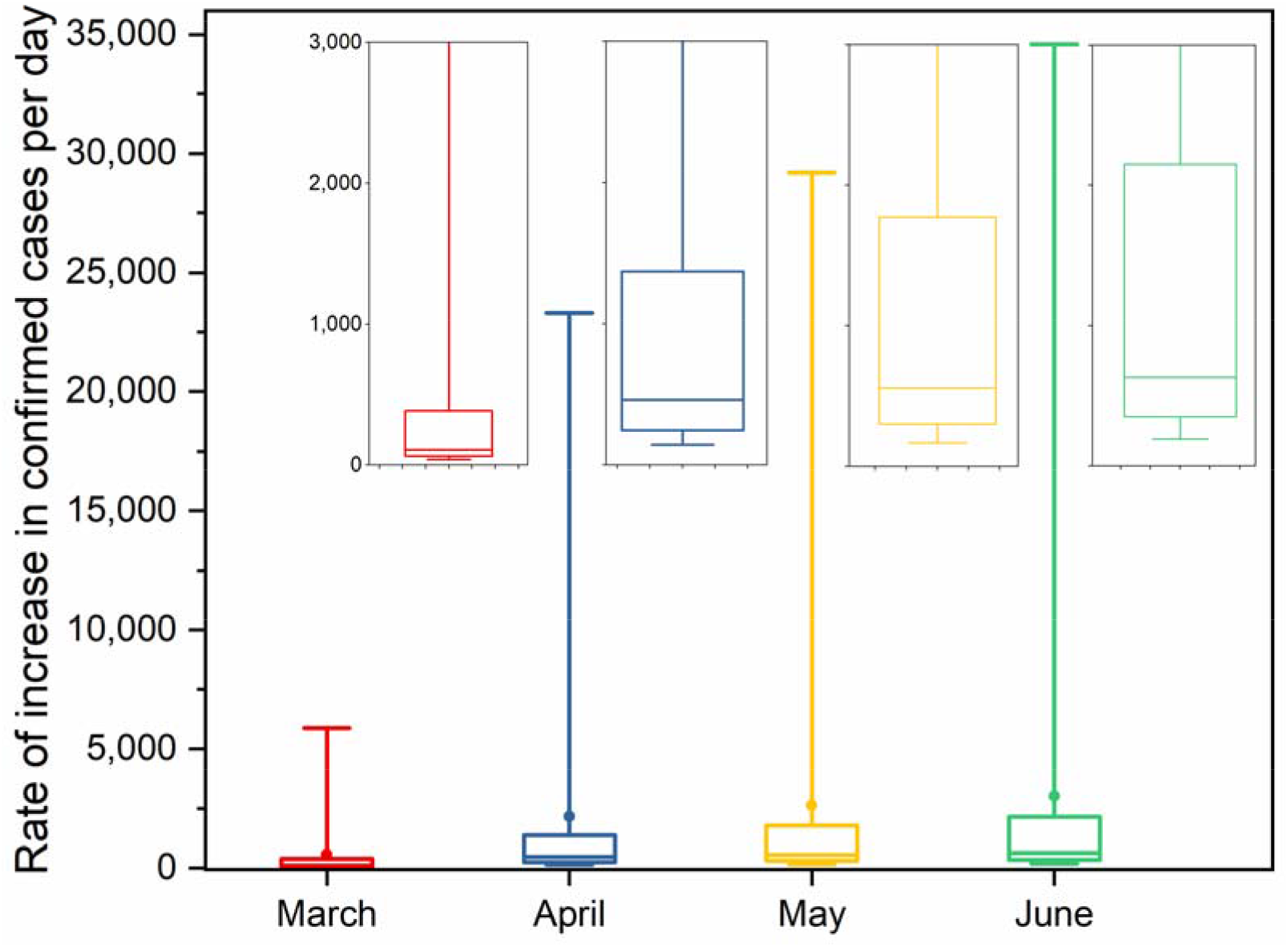
Rate of increase in confirmed cases per day (box plot illustrates variation in the rate from the data of 42 countries)

**Fig 6.**
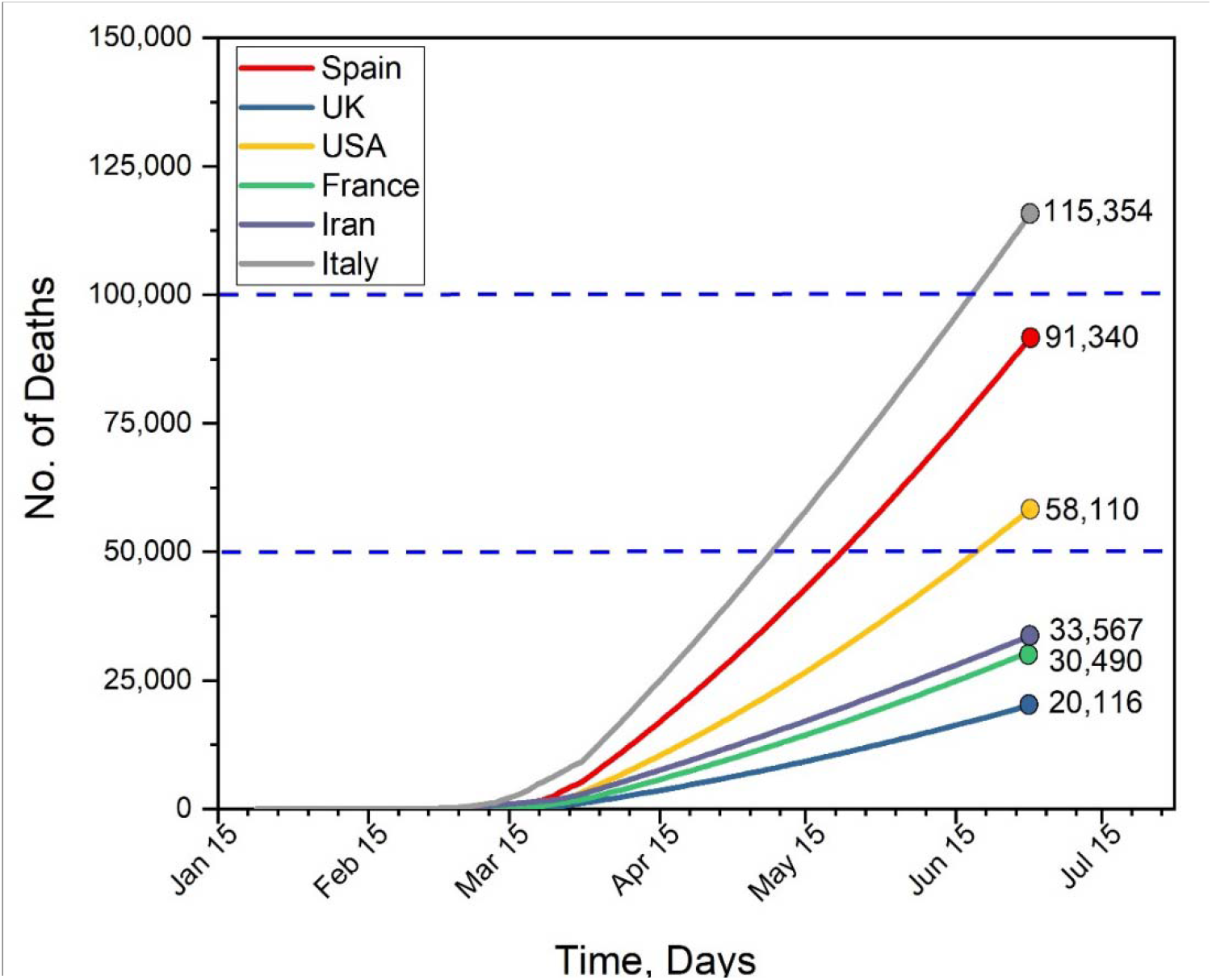
The estimated deaths in selected countries based the results of CFR and projection of confirmed cases.

**S5 Table: Estimated deaths for selected countries based on the projection of confirmed cases and average CFR value higher than 1% on June 30, 2020**

### Estimating fold increase due to the delay in reporting confirmed cases

Effective lag length is a key variable to estimate the fold increase due to delay in reporting. As different countries follow different testing procedure and also the capacity of health care systems largely varies between the countries, the results of confirmed cases on any day is lower than the actual number of people infected. We varied the minimum and maximum lag length of 1 and 7 days respectively to analyse the impact of delay on number of confirmed cases. Fig 7 is plotted between the number of days delayed and the number of folds increase in the confirmed cases for the selected countries across the world. The mean estimate of fold increase was calculated for each day lag (Fig 7). It is well known as illustrated in Fig 7 that increasing the number of days delay elevates the magnitude of fold increase. The fold increase of 16 and 10 would reach for the delay of 7 days in the context of extreme scenario as currently Italy and USA respectively are facing (Fig 7a). In specific, a steep increase was found when the lag length (i.e. delay in reporting) is more than 5 days. Therefore, it is to be noted that sooner the case is identified and reported, better the preventive measures could be ensured without much communal spread [30]. Further, we noticed that although USA is experiencing high surge in confirmed cases, the fold increase for even 1-week delay was considerably less compared to other countries. Perhaps this observation is likely to change as more data will be available. This is a positive point that USA can manage the situation if adequate preventive measures are in place. Overall, it was observed that many of the European countries exhibited similar results. As shown in Fig 7b, though the magnitude of fold increase seems comparatively low in less affected countries, it might increase when the number of confirmed cases increases. Therefore, this is a right time for them to enforce preventive measures to safeguard people from COVID-19.

**Fig 7.**
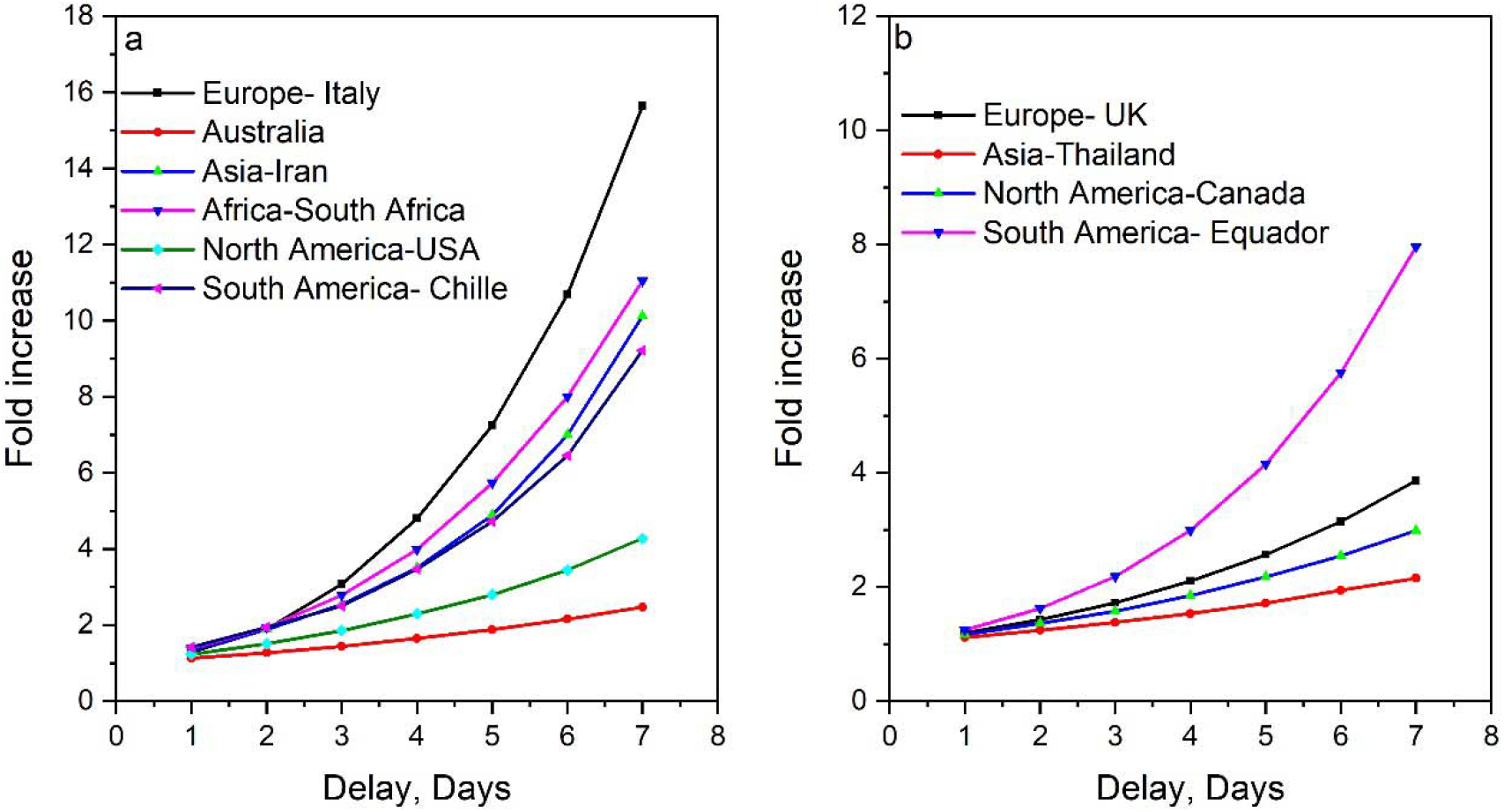
Fold increase in number of confirmed cases for different lag length a) severely affected b) less affected.

### Assumptions, limitations, and quantification of uncertainty

The uncertainties associated with the epidemic modeling studies are due to various reasons such as (i) availability, length, and correctness of the data [31], (ii) model parameter values: either assumed or estimated or adopted from previous modeling studies (assumption of incubation period and reproduction number) [32, 33] (iii) assumptions or limitations of the model being used. For example, the SEIR model assumes all the population is susceptible to infection. Dynamics transmission model’s assumption that symptomatic individuals are more (50%) susceptible to infection than asymptomatic individuals [32]. Assumptions while conceptualizing the non-pharmaceutical interventions such as duration of stay at home during isolation, percent contact reduction in workplaces, impact of non-pharmaceutical interventions are constant with time and same across all countries etc. [32].

As mentioned earlier, the uncertainty in the projected confirmed cases were quantified only based on the R_d_, and the fold increase estimated from each data point for a fixed lag length will have inherent variability (Fig. 8). This is mainly because the data is highly random and during the initial phase of pandemic the effect of delay will be less and gradually increase with time. Although the mean estimate as reported in the previous section is a good choice to quantify the delay effect in the projection, ignoring the uncertainty might under predict the likely estimate of future period. Therefore, we estimated 95% confidence interval from the estimate of fold increase. From the lag length period of 1 to 7 days, we considered on an average of 3- and 5-days lag based on the delay in reporting to estimate the range of variation in the projection especially in the confirmed cases. We chose Australia, randomly, to illustrate the impact of uncertainty in the projection (Fig 8). It is evident from Fig 8 that the median projected confirmed cases and uncertainty increases with time. For example, the median projected confirmed cases increased from 6,958 on March 30 2020 to 108,447 on 30 June 2020 for R_d_ of 3 days. Similarly, the width of uncertainty band for projected confirmed cases on 31 March 2020 was 6,596-7,593 (upper bound – lower bound) while it has increased to 102,816-118,357 on June 30 2020. Refer Table 2 for median and CI intervals for selected seven countries.

**Table 2.**
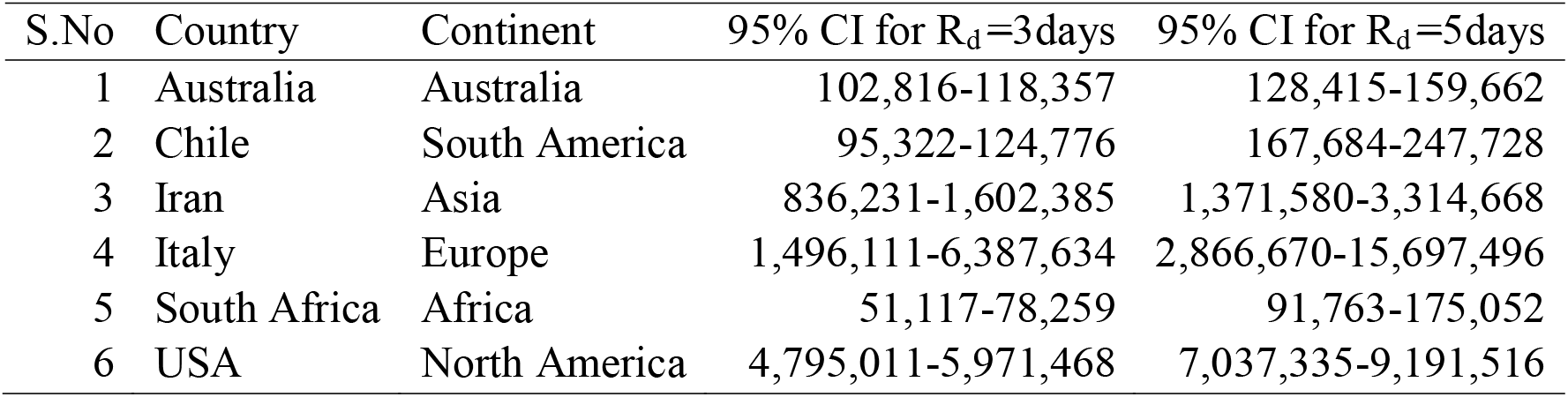
Projection of confirmed cases with uncertainty in Reporting delay of 3 and 5 days respectively for selected countries.

| S.No | Country | Continent | 95% CI for $R_d=3$days | 95% CI for $R_d=5$days |
| --- | --- | --- | --- | --- |
| 1 | Australia | Australia | 102,816-118,357 | 128,415-159,662 |
| 2 | Chile | South America | 95,322-124,776 | 167,684-247,728 |
| 3 | Iran | Asia | 836,231-1,602,385 | 1,371,580-3,314,668 |
| 4 | Italy | Europe | 1,496,111-6,387,634 | 2,866,670-15,697,496 |
| 5 | South Africa | Africa | 51,117-78,259 | 91,763-175,052 |
| 6 | USA | North America | 4,795,011-5,971,468 | 7,037,335-9,191,516 |

**Fig 8.**
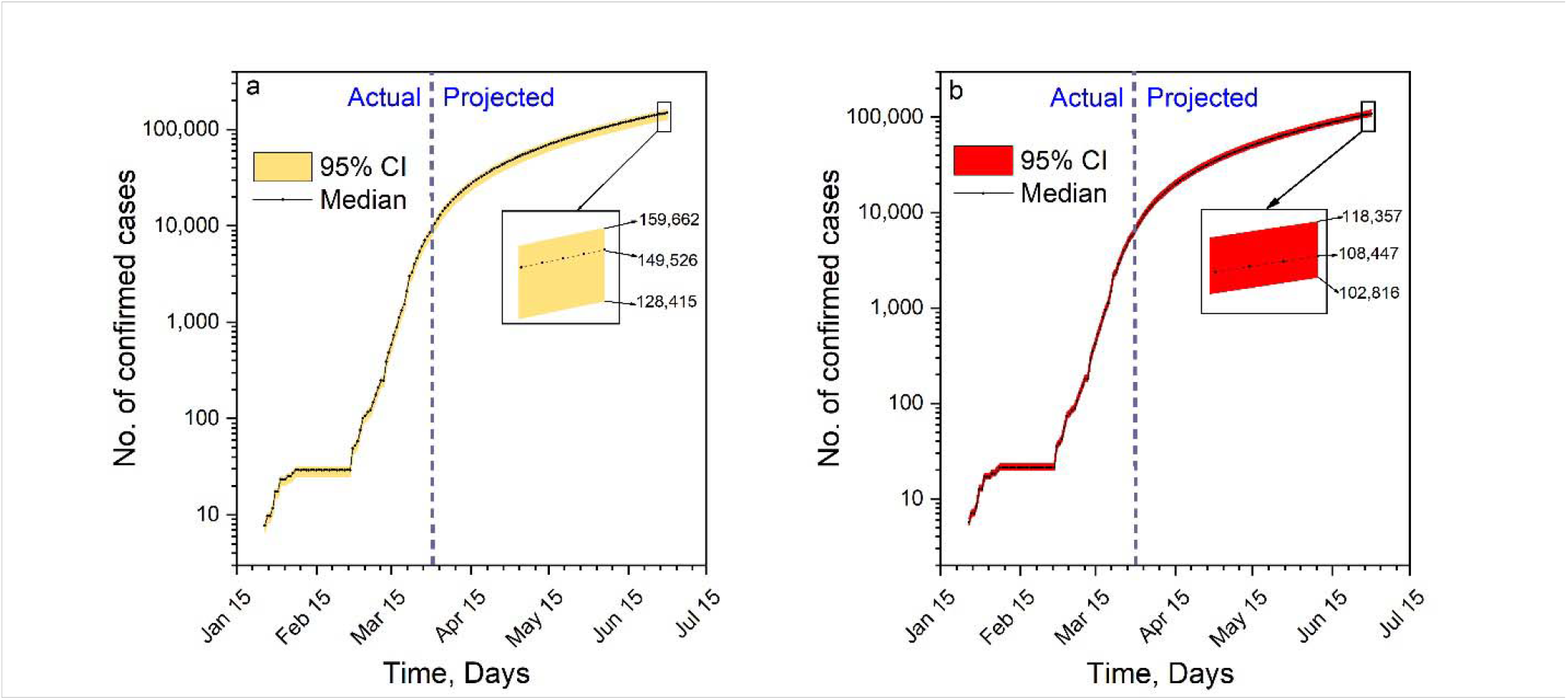
Actual and projected confirmed cases with uncertainty band for a) 5 days delay and b) 3 days delay for Australia.

It is expected that the unbiased estimate of uncertainty band will have the median estimate closer to the mid-portion of the band. In other words, any deviation from this mid-portion of the band represents the bias in the uncertainty estimate. However, we found the median falling towards the upper bound in most of the countries mainly due to the drastic increase in number of cases with during the critical period. As shown earlier in Fig 7, the delay in reporting increases the fold-increase in confirmed cases which in turn will significantly increase the projection range as well as the uncertainty (Table 2).

### Summary and limitations

This study explored the (i) applicability of EVDs in predicting the COVID-19 confirmed cases, and (ii) possible relation between the delay in reporting the cases and the potential increase in the number of infection (number of confirmed cases). The results of the projection indicate that the USA would have the highest number of confirmed cases of 2,905,522 (4,795,011-5,971,468 for R_d_= 3 days) and Iceland to have the minimum number of confirmed cases of 21,166 (27222-58,008 for R_d_= 3 days) by June 30, 2020. The number of deaths has also been estimated for the 42 countries and found that the deaths to be maximum in Italy (115,354 deaths) followed by Spain (91,340) by June 30, 2020.

It may be noted that we have not considered any intervention measures (i.e. lockdown, social distancing, school closures etc.,) in the model rather we focused on projecting the actual trend exist in the data, thereby informing the likely increase in number of confirmed cases due to COVID-19 outbreak. As inferred from this study, the reporting delay should be minimized to get more accurate information on the confirmed cases. Therefore, the uncertainty due to delay in reporting should not be ignored in the projection in order to estimate the reliable number of confirmed cases. However, future studies will include other sources of uncertainty such as model, parameter and input for the more realistic projection. The projected confirmed cases are based on the data collected until March 30, 2020. However, we will be updating the model projection once in every two weeks and our results will be posted in twitter handle @Hydroviswa and @ IdhayaI.

## Data Availability

The data were retrieved by the Center for Systems Science and Engineering(CSSE) at Johns Hopkins University: https://github.com/CSSEGISandData/COVID-19 [accessed on March 31, 2020].

## Ethics approval and consent to participate

The ethical approval or individual consent was not applicable

## Acknowledgement

Authors would like to thank Editor and anonymous reviewers for reviewing the paper. All authors would like to sincerely acknowledge their family members for supporting and encouraging to carry out the research work during this critical lockdown period.

## Authors Contribution

Conceived and designed the experiments: KSK. Performed the experiments: MA, KSK. Analyzed the data: KSK, MA, II, BS. Contributed reagents/materials/analysis tools: MA, MB. Wrote the paper: KSK, II, MA, BS. Interpretation of results: KSK, II, MA, JH, MB. Developed the codes: JH, KSK, MA.

